# Geographical characteristics and influencing factors of influenza epidemic in Hubei, China from 2009 to 2019

**DOI:** 10.1101/2023.01.05.23284238

**Authors:** Yang Mengmeng, Gong Shengsheng, Huang Shuqiong, Huo Xixiang, Wang Wuwei

## Abstract

Influenza is an acute respiratory infectious disease to which the population is generally susceptible and has an important impact on public health. Time series analysis and geospatial analysis were used to analyze the spatial and temporal distribution characteristics of influenza epidemic and its influencing factors in 103 counties in Hubei Province from 2009 to 2019, and the results showed that: 1) Influenza in Hubei Province mostly occurs in winter and spring, and the influenza epidemic is high from December to March every year, and there is an aggravating trend of influenza epidemic in recent years. 2) There were significant spatial and urban-rural differences in influenza prevalence in Hubei Province, with the eastern region being more serious than the central and western region, and the urban region more serious than the rural region. The hot spot areas of interannual variation of influenza were mainly distributed in the east and west of Hubei province, and the cold spot areas were distributed in the north-central of Hubei province. In addition, the cold hot spot areas of influenza epidemic varied from season to season. 3) The seasonal variation of influenza epidemic in Hubei Province is mainly governed by meteorological factors such as temperature, sunshine, precipitation, humidity, wind speed, etc. Low temperature, less rain, less sunshine, low wind speed and humid weather will increase the probability of influenza; the interannual variation and spatial variation of influenza epidemic is mainly influenced by socio-economic factors such as road density, number of beds per 1,000 people, urbanization rate and population density, and the intensity of the effect of these factors on influenza incidence rate in Hubei Province has significant spatial variation, but in general, the formation of spatial variation of influenza in Hubei Province is still the result of the joint action of socio-economic factors and natural meteorological factors.

## 1. Introduction

Influenza is an acute respiratory infectious disease caused by influenza virus, which mainly through by air, droplet and contact transmission. And influenza has a short incubation period, highly contagious, the virus is susceptible to mutation, the population is generally susceptible, the incidence rate is high, and the mortality rate is higher during the pandemic. Since the 20th century, there have been several influenza pandemics around the world, such as the “Spanish Flu” in 1918, the “Asian Flu” in 1957, the “Hong Kong Flu” in 1968, and the “Influenza A” in 2009, all of which were caused by the mutation of the Influenza A virus into a new type of virus, and some studies believe that there will be an influenza pandemic in the future, which will pose a major threat to human health. Before 2015, the number of reported influenza cases in China accounted for 3.05% of the total number of infectious disease cases, which has been climbing in recent years, and the proportion increased to 9.61% in 2021, and the prevention and control of influenza disease should not be relaxed in today’s world where new respiratory infectious diseases such as SARS, H1N1 and COVID-19 are constantly emerging.

Regarding the spatial and temporal distribution characteristics of influenza, previous studies suggest that influenza epidemics have obvious spatial and temporal aggregation, with temperate regions mostly occurring in winter and spring, while tropical and subtropical regions have two peak periods of influenza incidence in winter, spring and summer^[1]^.Regarding the influencing factors of influenza, previous studies have shown that meteorological factors such as temperature, humidity, sunshine, wind speed, precipitation, and atmospheric particulate concentration have a significant impact on the survival and spread of influenza viruses, and influenza viruses attached to atmospheric particulate matter have a higher chance of being exposed to and inhaled by the population^[2-3]^. In temperate regions, influenza viruses are mostly spread in the form of aerosols, and the low temperature, low humidity and low radiation in winter are favorable to the survival of influenza viruses, while in subtropical and tropical regions, the opposite is true, and influenza is mainly prevalent in the rainy season with high temperature and high humidity^[4-7]^. Crowding and population movement have an important influence on the susceptibility of the population to influenza, and closed clustered places such as schools, childcare institutions, and air travel are conducive to influenza epidemics^[8-10]^; influenza vaccination is the most effective means of preventing influenza viruses. Once the new influenza virus, suitable weather factors, and susceptible people appear at the same time, it is easy to cause an influenza outbreak epidemic. In general, medical scholars focus on influenza epidemiology, virology and immunology, while geographers focus on influenza spatial and temporal patterns, influencing factors and simulation prediction.

The population characteristics and seasonal characteristics of influenza epidemics in Hubei Province have been studied^[11-13]^, but its spatial characteristics and influencing factors are not yet studied. Hence, from the perspective of health geography, this study uses time series analysis and geospatial analysis to research the spatial and temporal distribution characteristics and influencing factors of influenza epidemics in Hubei Province from 2009 to 2019, with the aim of providing a scientific basis for the long-term prevention and control of influenza in Hubei Province.

## 2. Data sources and Research methods

### 2.1 Data sources

#### 1) Influenza incidence data

The number of influenza incidence in 103 counties in Hubei Province from 2009 to 2019 was obtained from the Hubei Disease Prevention and Control Information System (http://www.hbcdc.com/), and the incidence rate was calculated based on the number of incidence and population. Based on the purpose of the study and data availability, provincial-scale influenza data from 2009 to 2019 were used to analyze temporal changes of influenza epidemics in Hubei Province; county-scale influenza data from 2009 to 2019 were used to analyze the spatial differentiation of influenza epidemics in Hubei Province; When county-scale average incidence data from 2017 to 2019 were used to analyze the influencing factors of influenza epidemic in Hubei Province, this was because, Hubei has entered a high influenza season since 2017.

#### 2) Influencing factor data

Infectious disease epidemics are the result of a combination of three factors: pathogens, transmission routes and susceptible populations. First, regarding the pathogen factors, the survival and reproduction of influenza virus and its transmission are mainly governed by climatic conditions, so the annual average temperature (*x*_1_), annual average relative humidity (*x*_2_), annual average precipitation (*x*_3_), annual average wind speed (*x*_4_), and annual sunshine hours (*x*_5_) were selected for analysis in this study. 2017-2019 meteorological data were obtained from the China Meteorological Data website (http://data.cma.cn/). Secondly, regarding the transmission route factors, influenza is mainly transmitted by air, droplet and contact, and is closely related to population distribution and population mobility, so population density (*x*_6_), urbanization rate (*x*_7_), and road density (*x*_8_) are selected for analysis in this study. Finally, regarding the susceptible population factors, the population is generally susceptible to influenza, especially children and adolescents, and schools are the places where children and adolescents are concentrated and where influenza is highly prevalent; the immunity of the population to influenza is mostly limited by the regional health care level, the education level of the population and the economic conditions of the family, so the number of beds per 1,000 people (*x*_9_), the number of schools (*x*_10_) and the disposable income per capita (*x*_11_) are selected in this study for analysis. Among the study, the socioeconomic data from 2017 to 2019 were obtained from various statistical yearbooks of Hubei Province and China County Statistical Yearbook.

### 2.2 Research methods

#### 2.2.1 Influenza spatial and temporal characterization methods

##### Time series seasonal decomposition model

Based on the confirmation of the existence of seasonality of influenza incidence, a seasonal decomposition model was used to analyze the seasonality and long-term trend of influenza incidence in Hubei Province. Infectious disease time series are generally composed of four elements: Long-term trend (*T*), Seasonal variation (*S*), Cyclical variation (*C*) and Random variation (*R*). In general, the commonly used methods are additive and multiplicative models, which are used if the magnitude, trend and period of seasonal variation in incidence varies over time, otherwise, additive models are used. Referring to existing studies^[14]^ and the characteristics of the time series of influenza incidence, this study uses a multiplicative model (*X*_*i*_ = *T* × *C* × *S* × *R*) to decompose the month-by-month influenza incidence rate. The specific steps are as follows:

First, the moving average *MA*_*i*_ of the 12-month influenza incidence rate was calculated as follows.

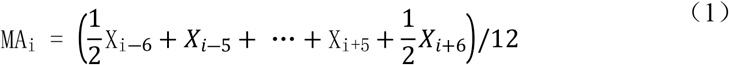

where *X*_*i*_ denotes the influenza incidence rate in month *i*. Cyclic variation and long-term trend series were obtained by moving average to eliminate seasonal variation and random variation, hence, *MA*_*i*_= *T*×*C*.

Second, the seasonal irregularity component *SR*_*i*_, which contains both seasonal and stochastic factors, was computed as follows.

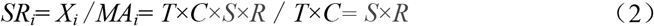

Then, *SR*_*i*_ was decomposed into seasonal factors *S* by eliminating the randomness factor *R* with monthly averaging, and normalized to obtain the average state and pattern of influenza incidence over time.

Finally, the *MA*_*i*_ serial values were curve-fitted to obtain the equation of long-term trend *T* of influenza incidence rate, aimed to reflect the overall fluctuation pattern of influenza incidence rate.

##### Spatial autocorrelation analysis

Spatial autocorrelation analysis is mainly used to check whether the attribute values of spatial units are correlated with those of their neighboring units, and is an important method to measure the degree of clustering of the attribute values of spatial units, which is divided into global spatial autocorrelation analysis and local spatial autocorrelation analysis. In this study, global spatial autocorrelation is used to analyze the overall characteristics of the spatial distribution of influenza in Hubei Province, which is measured by Moran’s *I*. Moran’s *I* > 0 indicates the existence of positive spatial correlation, which is an agglomerative distribution, and Moran’s *I* < 0 indicates the existence of negative spatial correlation, which is a discrete distribution. Then Getis-Ord *Gi\** local spatial autocorrelation was used to analyze the local characteristics of the spatial distribution of influenza in Hubei Province, which was measured by *Gi\** index, *Gi\** > 0 indicates the existence of hot spot area, and *Gi\** < 0 indicates the existence of cold spot area. This analysis process was performed in Arc GIS software.

#### 2.2.2 Methods for analyzing influencing factors of influenza

##### Geographical Detector

Geographic detector(GD) can effectively detect the spatial heterogeneity of geographic phenomena and their drivers^[15]^. In this paper, single factor detection and interaction detection are used to identify the factors influencing the spatial differentiation of influenza incidence rate in Hubei Province, which are calculated as follows:

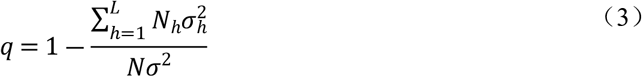

Where, the value of *q* is the decisive indicator to measure the spatial variation of influenza incidence, the value range of *q* is [0,1], the larger the value of *q*, the greater the influence of the factor on the spatial variation of influenza incidence; *h*=1,……, *L* is the number of categorical sub-regions of the influencing factor; *N*_*h*_ is the sub-region *h, N* is the number of spatial units, here *N*=103, indicates 103 counties in Hubei province; 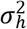 and *σ*^2^ are the discrete variances of the influenza rate in sub-region *h* and Hubei province, respectively.

##### Geographically weighted regression analysis

Geographically weighted regression model (GWR) is a spatial regression method based on the idea of local smoothness, which can be used to analyze the spatial variability of the degree of different influencing factors on the influenza rate. This method takes into account the geospatial attributes of the data into the model and is an extension of the traditional multiple regression model, focusing on the variability and variability of the regression coefficients within geographic space^[16]^. The model is expressed as follows:

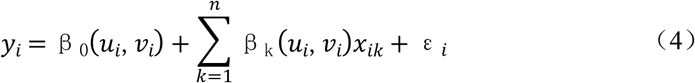

Where, *y*_*i*_ is the dependent variable of the sample, which is the influenza rate of a district or a county; *x*_*ik*_ is the matrix of influencing factors; (*u*_*i*_,*v*_*i*_) is the spatial location coordinates of sample *i*;*β*_*k*_ (*u*_*i*_,*v*_*i*_) is the regression coefficient of influencing factor *k*; *ε*_*i*_ is the random error term. In this study, geographically weighted regression analysis of influencing factors of influenza rate was performed with the help of ArcGIS software, and the kernel type was set as fixed distance, and the bandwidth method was the minimum AIC method.

## 3. Results

### 3.1 Temporal changes of influenza epidemic in Hubei Province

#### 3.1.1 Seasonal variation characteristics of influenza

From 2009 to 2019, the average monthly number of influenza cases in Hubei Province was 3187, and the average monthly incidence rate was 5.40/100,000, with a significantly higher number/rate of influenza incidence between December-sub-March (Fig 1). After time series analysis, the seasonal component *S* of influenza incidence rate in Hubei province was obtained to form a peak value in December-sub-March, indicating that influenza in Hubei province is mainly prevalent in winter. According to the previous study, the peak period of influenza in southern China is summer, and the peak period of influenza in northern China is winter and spring^[17]^. Hubei province is located in the south but adjacent to the north, and the influenza epidemic has certain transitional characteristics between the north and the south; so, in addition to the peak epidemic period in winter, there is also a small high influenza period in August in summer (Fig 2).

**Fig. 1.**
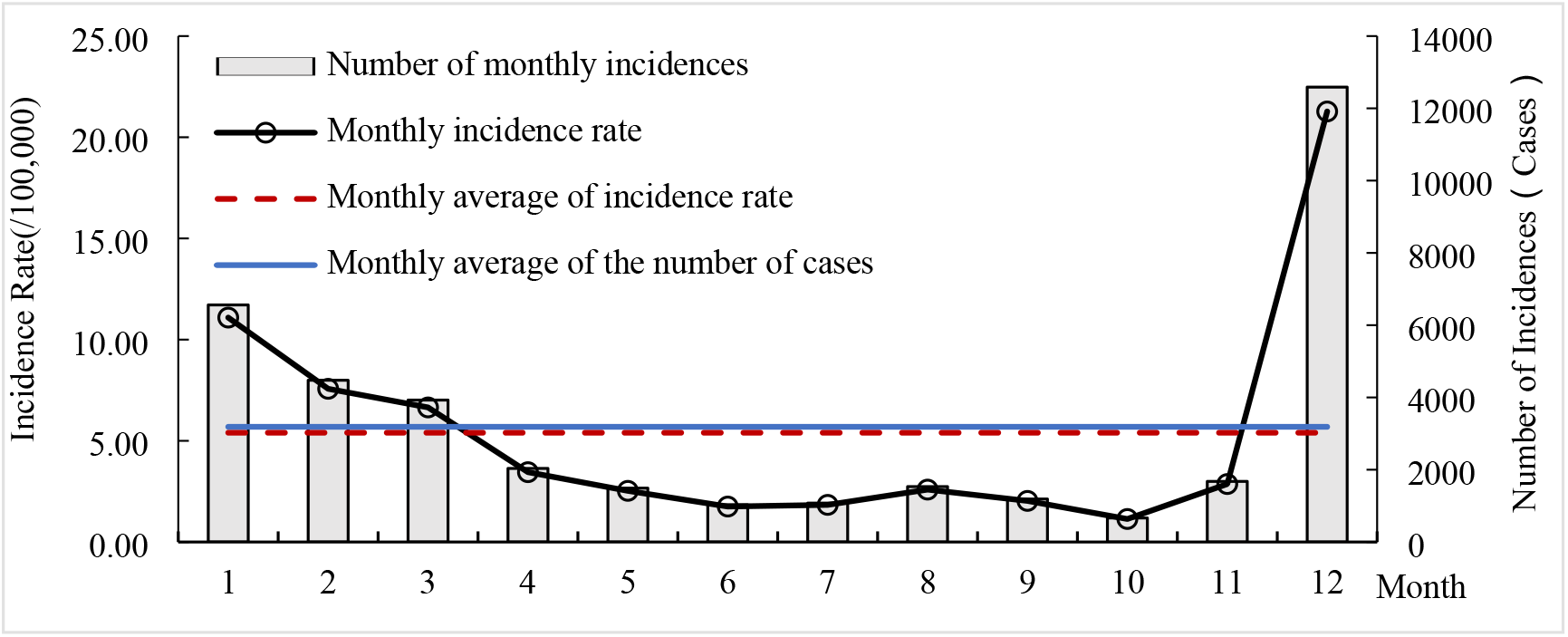
Monthly changes of cases and incidence rate of influenza in Hubei during 2009-2019

**Fig. 2.**
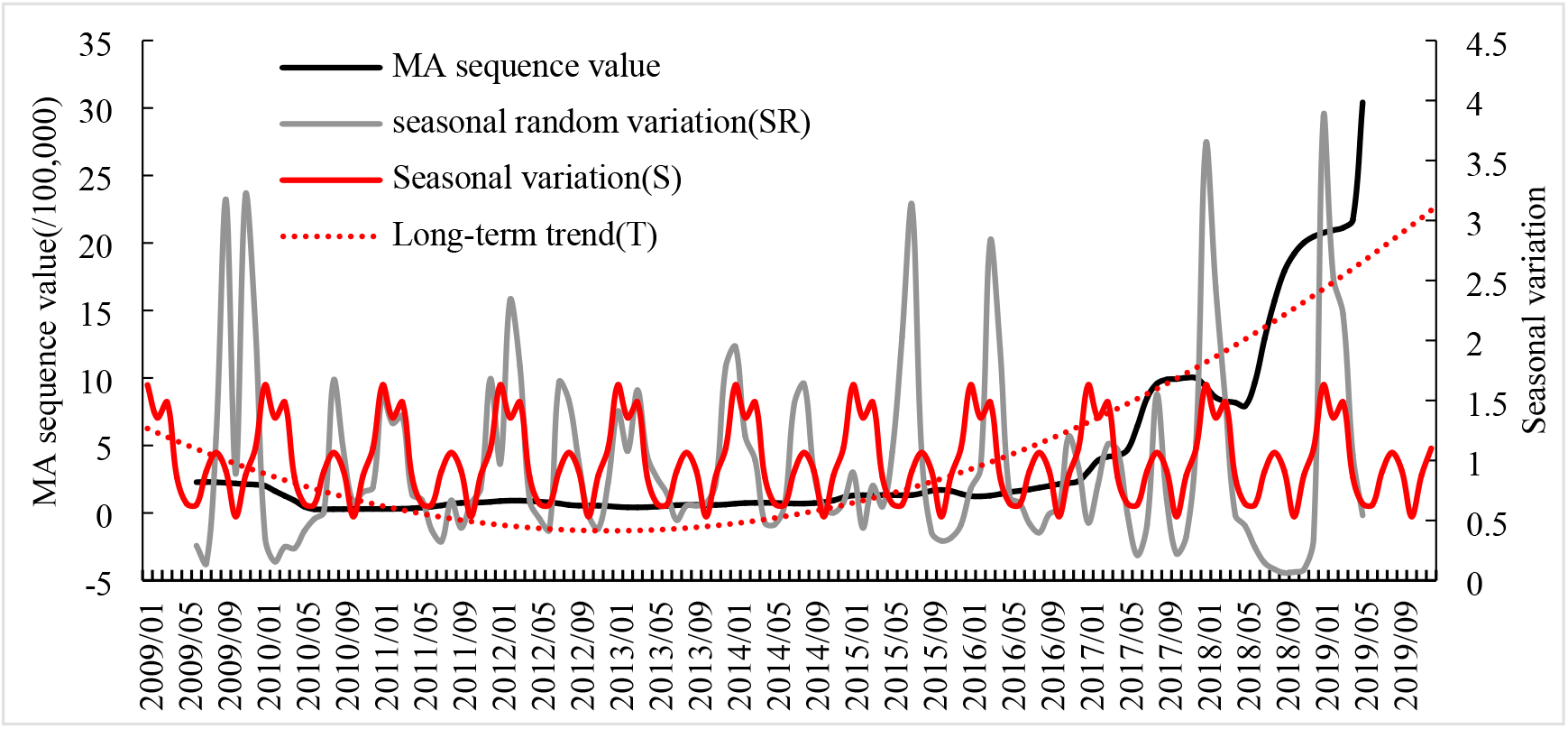
Seasonal index and long-term trend of influenza incidence in Hubei during 2009-2019

#### 3.1.2 Interannual variation characteristics of influenza

From 2009 to 2019, the number of influenza cases/incidence rate in Hubei Province decreased and then increased (Fig 3), with the number of cases decreasing from 15,444 in 2009 to 1994 in 2010 and then increasing to 274,611 in 2019, and the incidence rate decreasing from 27.04/100,000 in 2009 to 3.49/1,000 in 2010 and then increasing to 463.31/100,000 in 2019. Both the number of cases and incidence rate in 2019 were the highest in the study period, but there was only a winter peak and no summer peak in that year (Fig 2), which was more concentrated and could be considered a pandemic year for influenza. The reasons for this phenomenon may be related to the absence of two summer peaks in 2018 and 2019, and the occurrence of antigenic mutations of influenza A virus^[11]^.

**Fig. 3.**
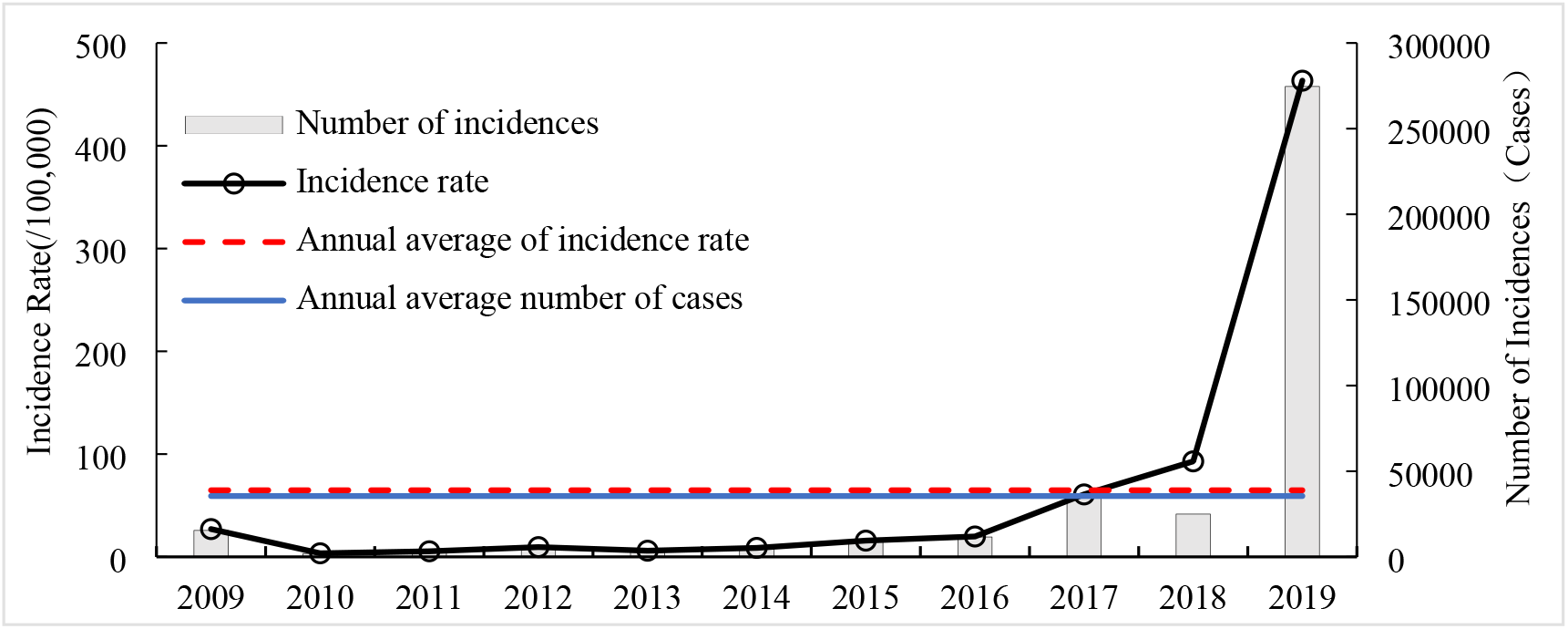
Interannual changes of cases and incidence rate of influenza in Hubei during 2009-2019

#### 3.1.3 Characteristics of long-term trends in influenza

After analyzing the time series of influenza incidence rate in Hubei Province from 2009 to 2019 (Fig 2), the long-term trend equation can be obtained as *T* = 0.0034*t*^*2*^ - 0.3272*t* + 6.5793 (*R*^2^ = 0.8416). From this quadratic polynomial equation, it can be seen that the overall change of influenza rate in Hubei Province from 2009 to 2019 showed a decline followed by an increase, and there was an aggravating trend in recent years.

### 3.2 Spatial distribution of influenza epidemic in Hubei Province

#### 3.2.1 Overall distribution characteristics of influenza

1) Influenza is widespread, but the distribution varies significantly. In terms of the number of cases (Fig 4a), from 2009 to 2019, influenza in Hubei Province was mainly distributed along the “A-type urban skeleton”^[18]^, with Ezhou, Wuhan, Huanggang, Huangshi, Jingzhou, Yichang, and Shiyan along the route being highly concentrated areas of influenza cases. By region, the proportion of national land area in the east, middle and west of Hubei province was 21.73%, 36.19% and 42.36%, respectively, and the corresponding cumulative “concentration” of influenza cases (the ratio of the proportion of influenza cases to the proportion of national land area) was 2.11, 0.81 and 0.59, respectively. The distribution of influenza cases in the eastern, central and western longitudinal directions is very obvious. By urban and rural areas, the 64 main urban areas and county-level cities are roughly classified as “urban areas” and the remaining districts and counties are classified as “rural areas”, and the cumulative “concentration” of influenza cases corresponding to these two types of areas are 1.54 and 0.50 respectively, with significantly more cases in urban areas than in rural areas. In summary, the overall distribution of influenza epidemic in Hubei Province is characterized by: eastern areas are more serious than central and western areas, and urban areas are more serious than rural areas.

**Fig. 4.**
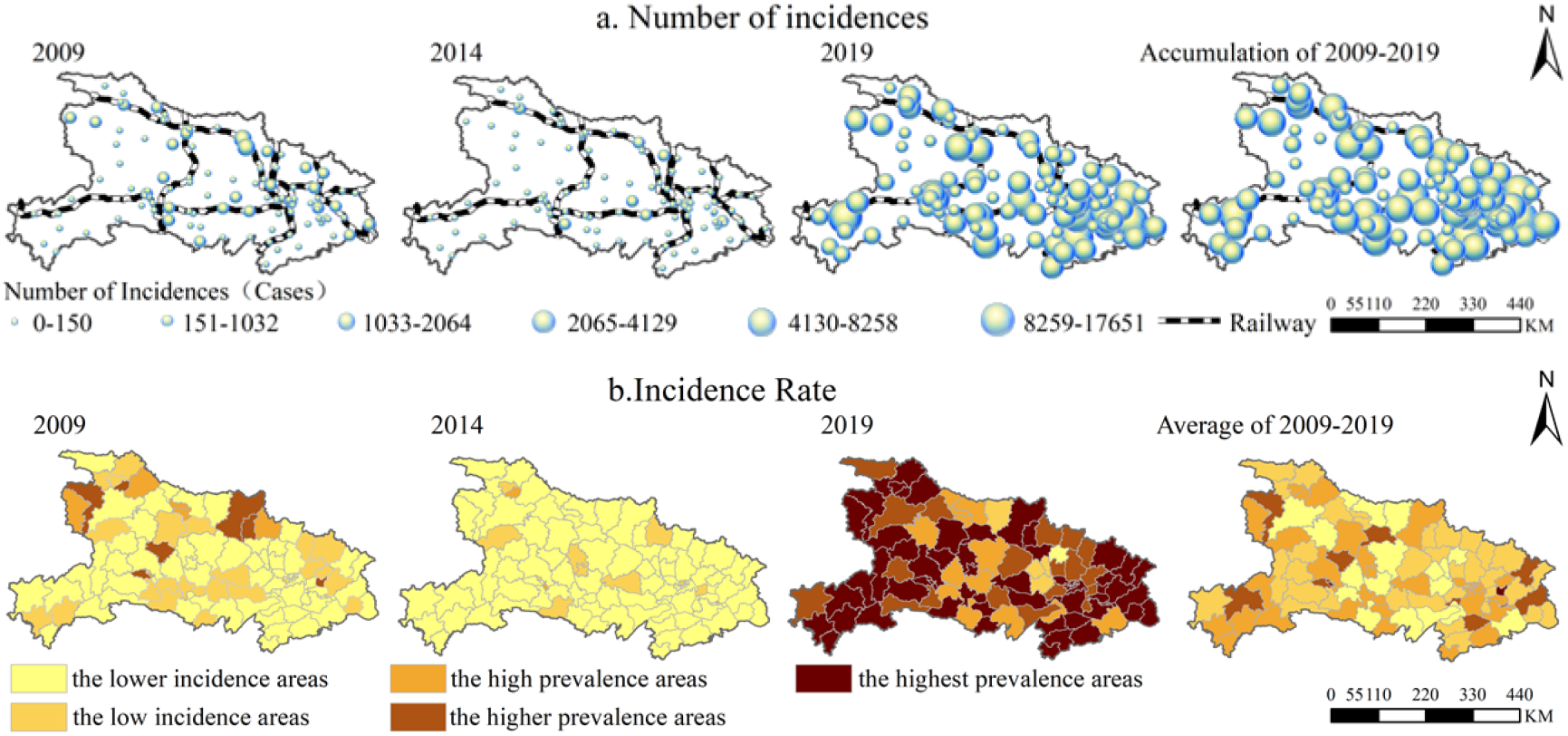
Distribution of influenza cases (a) and incidence rate (b) in the country scale in Hubei during 2009-2019

2) The scope of the high incidence rate area of influenza gradually expanded, and the scope of the low incidence rate area first expanded and then shrinked. In terms of incidence rate, the county influenza incidence rate in Hubei province from 2009 to 2019 was divided into 5 levels according to the natural breakpoint method, and the 5 levels of areas were named with reference to the average annual influenza incidence rate in Hubei province (64.81/100,000): the lower incidence areas (<22.60/100,000), the low incidence areas (22.60/100,000-64.81/100,000), the high prevalence areas (64.81/100,000-129.62/100,000), the higher prevalence areas (129.62/100,000-259.24/100,000), and the highest prevalence areas (>259.24/100,000) (Fig 4b). Among them, in 2009, the “the higher prevalence areas “ were scattered in Maohuai, Zhushan, Zengdu, Suixian, Doujun, Yuan’an, Huangzhou and other counties, with the number of counties accounting for only 6.8%, while in 2019, all the province was covered with “ the highest prevalence areas”, with the number of counties accounting for 63.11%, mainly located in Huanggang, Xianning, Wuhan, Shiyan, Enshi, Yichang and other cities in the east and west of Hubei province. From 2009 to 2019, the number of counties in “ the lower incidence areas “ accounted for 63.11%, 87.38% and 0.97% respectively, with a trend of spatial distribution expanding first and then narrowing. It can be seen that the spatial distribution of influenza epidemic in Hubei Province in recent years has a clear trend of spreading, which is consistent with the previous conclusion of temporal changes.

#### 3.2.2 Spatial clustering characteristics of influenza

1)Influenza showed an overall clustered distribution, with the strongest clustering in winter. In terms of interannual distribution, the influenza epidemic in Hubei Province showed a clustered distribution during the study period except for 2014-2015, with the strongest clustering in 2013 (*I* = 0.32, *P* < 0.001). In terms of seasonal distribution, 2017.01, 2017.07-10, 2017.12, 2018.03-04, 2019.03-06, and 2019.09-12 all showed a clustered distribution, with the strongest clustering in 2019.12 (*I* = 0.48, *P* < 0.001). In short, the influenza epidemic in Hubei Province showed a clustered distribution, with the strongest degree of clustering in winter.

2) Influenza hot spot areas were located in the east and west of Hubei Province, and cold spot areas were located in the north-central of Hubei Province. In terms of interannual changes (Fig 5a), the hot spot areas migrated to the southeast and the cold spot areas moved slightly to the north. In 2009, the hot spot areas were distributed in the west of Hubei province, with 9 counties and districts, and the cold spot areas were distributed in the suburbs of Wuhan. In 2014, the hot spot areas were distributed in 14 counties and districts in the northwest and southwest of Hubei province, and there was no cold spot areas. In 2019, the hot spot areas in the southwest of Hubei province still existed, and the hot spot areas in the southeast of Hubei province was added, with 21 counties and districts, and the cold spot areas were distributed in the north-central of Hubei province, with 6 counties and districts. For the whole study period of 2009-2019, there are two hot spot areas of influenza epidemic in west and east of Hubei province, and the cold spot areas are mainly distributed in the north-central of Hubei province. In terms of seasonal changes (Fig 5b), the distribution of cold and hot spots areas of seasonal influenza epidemics in Hubei Province in 2017-2019 was similar to the interannual distribution, with hot spot areas in spring and autumn mainly distributed in the west of Hubei province, hot spot areas in summer mainly distributed in the southeast of Hubei province, and two hot spot areas in winter, southwest and southeast of Hubei province; cold spot areas have been stable in north-central of Hubei province. Influenza epidemic is often caused by a combination of factors, which is related to socio-economic factors such as population movement and concentration, and may also be related to the large temperature difference between winter and spring in the hilly areas of the west of Hubei province.

**Fig. 5.**
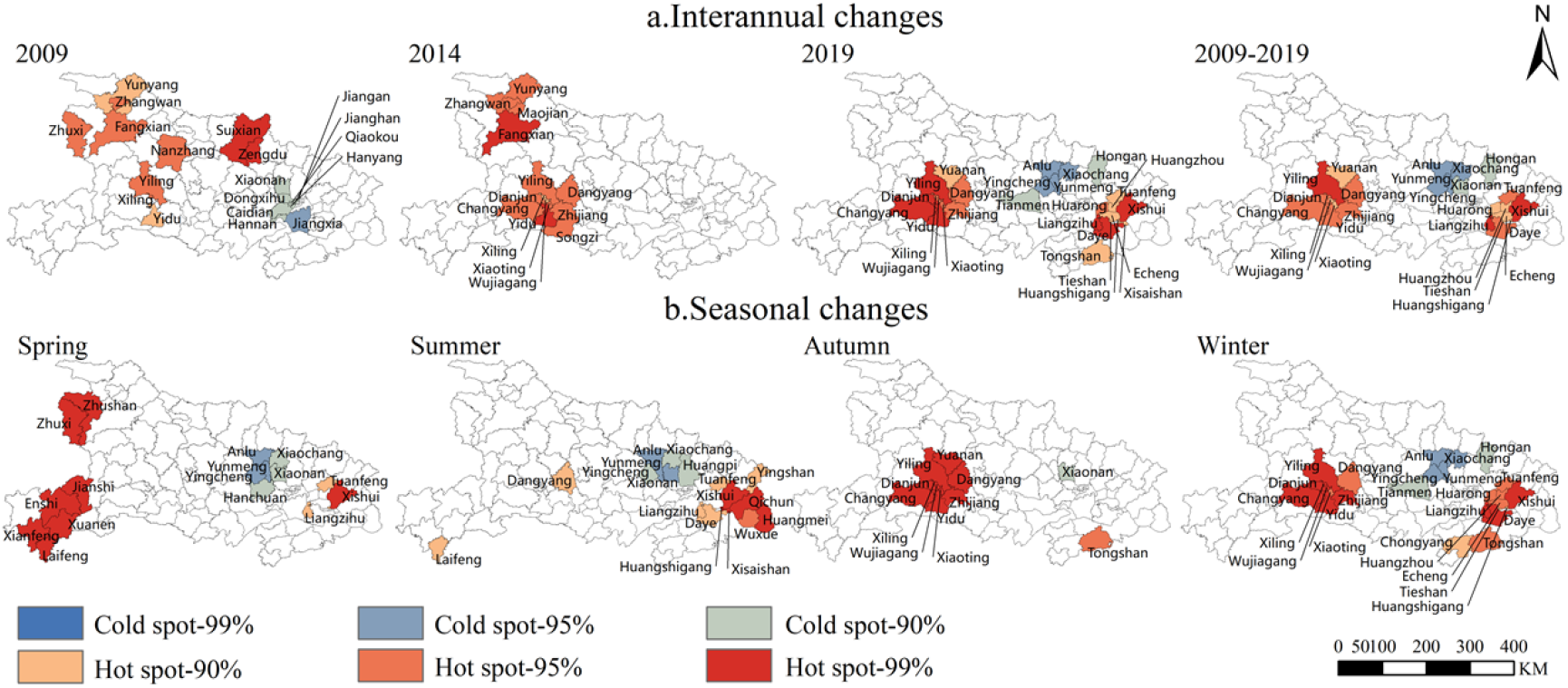
Interannual(a) and seasonal changes(b) of hot spots of influenza incidence in Hubei during 2009-2019

### 3.3 Influencing factors of influenza epidemic in Hubei Province

#### 3.3.1 Factors influencing the temporal changes of influenza

##### Factors influencing seasonal changes in influenza

Meteorological factors can affect the seasonal spread of influenza by influencing the survival rate of the virus outside the human body, transmission routes, population immunity, and behavioral habits^[19]^. Firstly, the monthly influenza incidence rates in Hubei Province from 2009 to 2019 were correlated with the monthly average temperature, precipitation, relative humidity, wind speed and sunshine hours during the same period, respectively, and the results showed that the overall influenza incidence rate was negatively correlated with the monthly average temperature (*r* = -0.19, *P* < 0.05) in a more significant way, indicating that influenza occurred more often in cold weather. Secondly, the influenza incidence rate of each district and county in Hubei Province from 2017 to 2019 was correlated with the monthly mean temperature, precipitation, relative humidity, wind speed, and sunshine hours of each district and county during the same period, and the results showed (Table 1) that the influenza incidence rate was correlated with the monthly mean temperature (*r*=-0.30), precipitation (*r*=-0.11), sunshine hours (*r*=-0.24), and wind speed (*r*=-0.07) all showed significant negative correlations (*P*<0.01) and significant positive correlations with relative humidity (*r*=0.08, *P*<0.01), indicating that influenza is more likely to occur in low temperature, less rain, less sunshine, low wind speed and humid weather. This is because low temperatures can prolong the survival time of influenza viruses in the environment^[20]^, especially in winter when cold weather drives people to increase their indoor activities, and lack of air circulation increases the probability of illness, as does cloudy weather, and conversely, long daylight hours and high UV intensity can effectively destroy and kill airborne viruses^[21]^; influenza viruses are more likely to combine with moisture in the air under conditions of high humidity and the increase in volume makes the rate of decline faster, combined with poor air mobility under low wind speed conditions, the virus can remain suspended in the air for a longer period of time^[22]^, thus increasing the probability of illness. In addition, the incidence rate of influenza is positively correlated with the average temperature in March, June, November and December, which indicates that sudden changes in temperature during seasonal transitions are more likely to trigger influenza epidemics, seasonal transitions and alternating heat and cold affect the regulation of the body’s thermal function to a certain extent^[23]^, leading to the occurrence and spread of influenza, especially in the elderly and frail. Influenza incidence was significantly negatively correlated with precipitation in March, April, and July, and significantly positively correlated with precipitation in June and August, indicating that the relationship between influenza and precipitation is more complex.

**Table 1.**
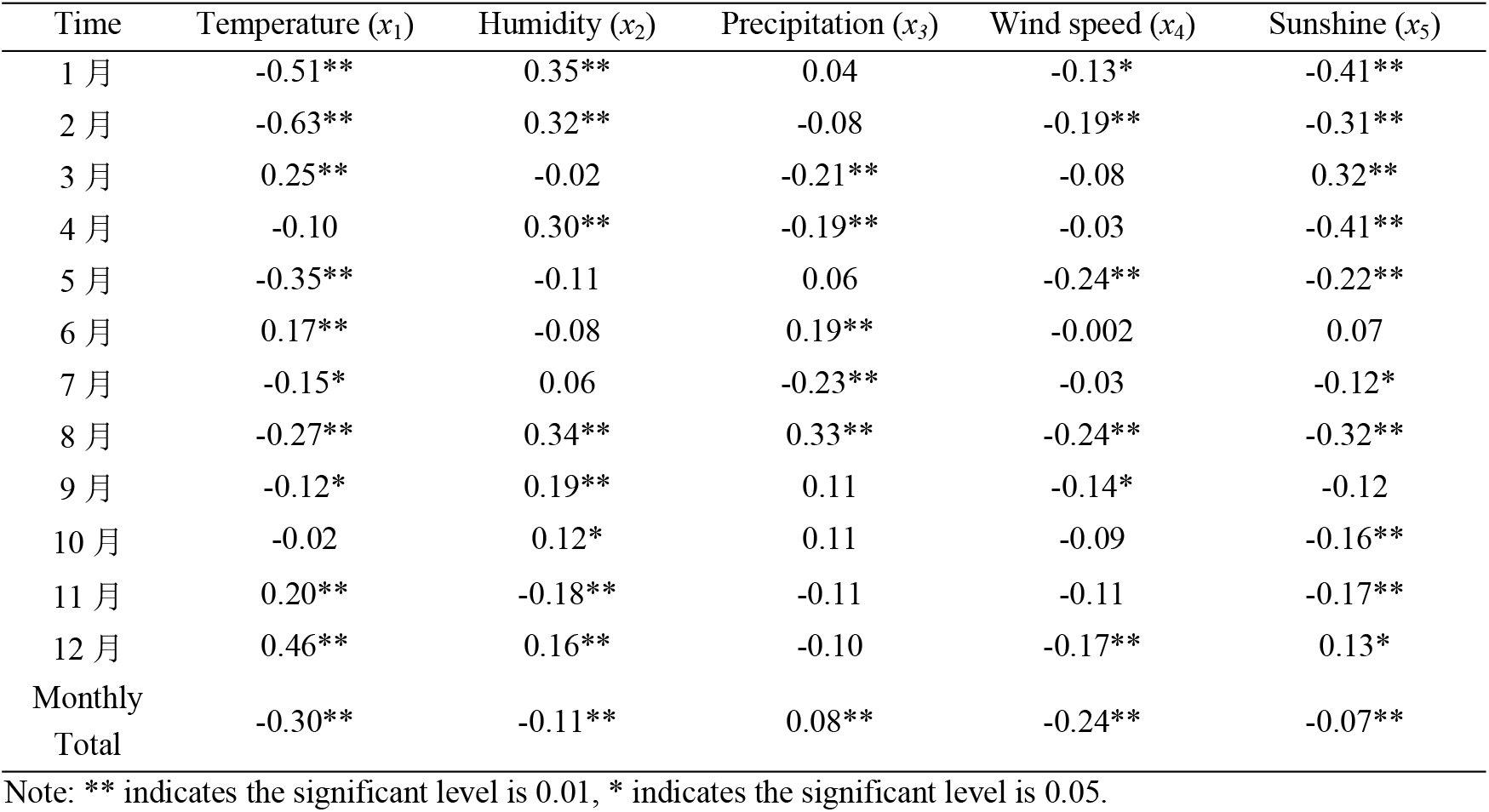
Spearman correlation analysis of influenza incidence in Hubei during 2017—2019

| Time | Temperature ($x_1$) | Humidity ($x_2$) | Precipitation ($x_3$) | Wind speed ($x_4$) | Sunshine ($x_5$) |
| --- | --- | --- | --- | --- | --- |
| 1 月 | -0.51** | 0.35** | 0.04 | -0.13* | -0.41** |
| 2 月 | -0.63** | 0.32** | -0.08 | -0.19** | -0.31** |
| 3 月 | 0.25** | -0.02 | -0.21** | -0.08 | 0.32** |
| 4 月 | -0.10 | 0.30** | -0.19** | -0.03 | -0.41** |
| 5 月 | -0.35** | -0.11 | 0.06 | -0.24** | -0.22** |
| 6 月 | 0.17** | -0.08 | 0.19** | -0.002 | 0.07 |
| 7 月 | -0.15* | 0.06 | -0.23** | -0.03 | -0.12* |
| 8 月 | -0.27** | 0.34** | 0.33** | -0.24** | -0.32** |
| 9 月 | -0.12* | 0.19** | 0.11 | -0.14* | -0.12 |
| 10 月 | -0.02 | 0.12* | 0.11 | -0.09 | -0.16** |
| 11 月 | 0.20** | -0.18** | -0.11 | -0.11 | -0.17** |
| 12 月 | 0.46** | 0.16** | -0.10 | -0.17** | 0.13* |
| Monthly Total | -0.30** | -0.11** | 0.08** | -0.24** | -0.07** |

Note: \*\* indicates the significant level is 0.01, \* indicates the significant level is 0.05.

##### Factors influencing interannual changes in influenza

Correlation analysis of the annual average influenza incidence rate of each district and county in Hubei Province from 2017 to 2019 with the annual average of 11 influencing factors showed that the influenza incidence rate was significantly and positively correlated (*P* < 0.01) with road density (*r* = 0.40), urbanization rate (*r* = 0.36), and number of beds per 1,000 people (*r* = 0.35), and with the number of schools (*r* = - 0.24) showed a more significant negative correlation (*P* < 0.05). A further stepwise regression of the 11 influencing factors showed that only road density (*r*=0.72), population density (*r*=-0.39), and sunshine (*r*=0.21) had a significant effect on influenza incidence (*y*=0.21*x*_*5*_-0.39*x*_*6*_+0.72*x*_*8*_). This all indicates that the main factors influencing the interannual changes of influenza incidence rate are socioeconomic factors, especially road density, meaning that they are mainly closely related to the local population mobility.

#### 3.3.2 Factors influencing the spatial differentiation of influenza

##### 1) The strength of the influencing factors

The results of detecting factors influencing the spatial changes of influenza incidence rate in Hubei Province using geographic detector showed that, in terms of single-factor detection, only three factors passed the *p* < 0.1 significance test, ranked by *q*-value: road density (0.22) > urbanization rate (0.18) > number of schools (0.12), indicating that socioeconomic factors are the dominant factors influencing the spatial changes of influenza incidence rate. In terms of interaction detection, the interactions of all types of factors strengthen the influenza epidemic, and the interactions between socioeconomic factors and natural meteorological factors are significantly greater than the interactions between socioeconomic factors and natural meteorological factors, such as the interactions between road density and precipitation (0.53), interactions between urbanization rate and number of schools (0.51), interactions between number of beds per 1,000 people and temperature (0.50), and interaction of road density and air temperature (0.49), and interaction of urbanization rate and humidity/wind speed (0.47) (Fig 6). Taken together, the formation of spatial changes in influenza incidence rate in Hubei Province is the result of the combined effect of socioeconomic and natural meteorological factors.

**Fig. 6.**
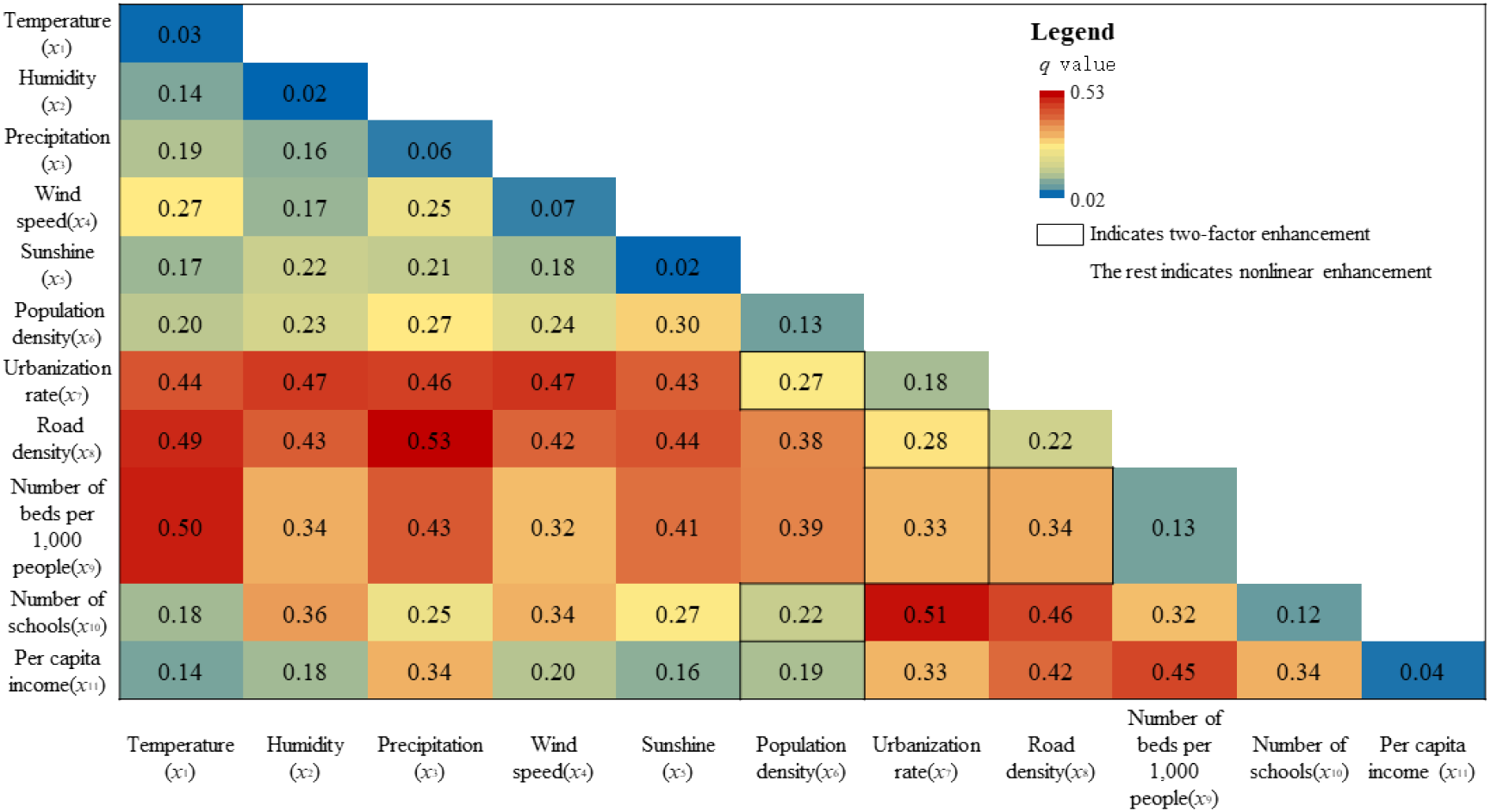
Interaction hotspot maps of influencing factors on influenza incidence in Hubei during 2017-2019

##### 2) Spatial variation in the degree of influencing factors

The results of the geographically weighted regression analysis showed that the effects of road density, number of beds per 1,000 people, urbanization rate, and population number on influenza incidence rate in Hubei Province had significant spatial differences (Fig 7), among which, the effect of road density and population density showed a positive correlation overall, with a gradient increasing trend from east to west, and the effect of road density was the greatest, indicating that the influence of population movement on influenza epidemic in west of Hubei province was significantly greater than that in central and east of Hubei province. The temperature difference is larger in the mountainous and hilly area in west of Hubei province, which affects the thermoregulatory function of human body, and the seasonal population inflow increases in winter and spring when a large number of students and workers return home, while the population outflow is mainly in east of Hubei province, so the effect on west of Hubei province is stronger. The effect of the number of beds per 1,000 people on influenza epidemic shows a positive correlation, with the strength of the effect being low in the central part and high on both sides, with the highest in the east of Hubei province; the number of beds per 1,000 people reflects the level of regional health care, and the stronger public health awareness and better consultation habits in the east of Hubei province, make the onset of influenza easier to be screened^[24]^. The intensity of the role of urbanization rate on influenza epidemic decreases from southeast to northwest; urbanization rate reflects the degree of population concentration, and the probability of influenza virus transmission is higher in urban areas with high population concentration^[25]^, and the urbanization level is higher in the east of Hubei province, and the intensity of the role of urbanization rate on influenza epidemic is also higher.

**Fig. 7.**
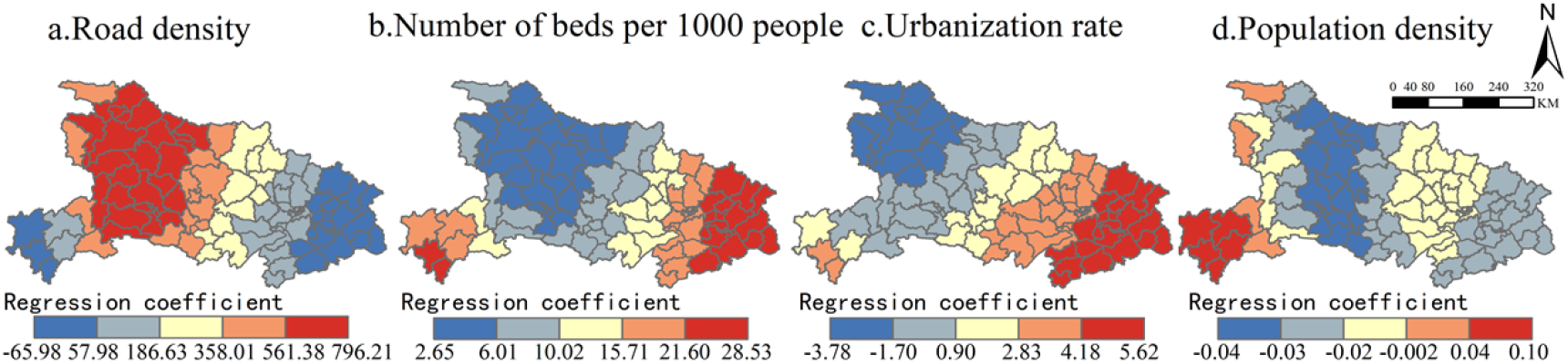
spatial distribution of regression coefficients influencing factors on influenza incidence in Hubei during 2017-2019

## 4. Discussion

1. The influenza epidemic in Hubei Province mainly peaks in winter during the year, which is similar to the epidemiological characteristics of Sichuan and Hunan, which are also mid-latitude regions^[26-27]^; in addition to the peak epidemic period in winter, there is also a small high incidence period in August in summer, which shows that the influenza epidemic in Hubei Province has certain north-south transition characteristics, which is similar to the epidemiological characteristics of Shanghai^[28]^. The number/rate of influenza incidence has increased rapidly since 2017, with 2019 being the highest recent year, similar to the situation in many domestic places during the same period^[29]^, which is related to the absence of two small high summer influenza periods in Hubei Province in 2018 and 2019, and the cyclical epidemic of influenza A virus (average 2-3 year mini-pandemic, 10-15 years pandemic) are related^[11]^. Therefore, influenza prevention and control efforts should focus on strengthening influenza vaccination in the winter and spring seasons, as well as daily cleaning and disinfection of key public places such as schools and amusement parks.
2. The clustering of influenza incidence cases in Hubei province is mainly in the eastern and urban areas, and the hotspot areas of incidence rate have a tendency to migrate from west to east, which has strong similarity with the epidemic of hand, foot and mouth disease in Hubei province^[30]^, and is highly consistent with the population migration trend in Hubei province in recent years^[31]^. Therefore, the priority areas for influenza prevention and control in Hubei province should be placed along the “A-type skeleton” cities with high urbanization levels, and the monitoring of migrant populations should be strengthened.
3. The low-temperature weather in Hubei Province is favorable for influenza epidemics and has similarities with influenza epidemics in Hunan Province, Suzhou City, and Beijing^[26-32-33]^, which reminds us to pay attention to influenza viruses during low-temperature weather in winter. In addition, further research in this study found that cloudy, overcast, or low wind speed weather conditions, where air circulation becomes poor, can prolong the survival time of influenza viruses and increase the chance of illness. Therefore, residents should focus on daily influenza prevention and control by diligent ventilation, less gathering, wearing masks and vaccination.
4. Influenza incidence rate in Hubei province was positively correlated with urbanization rate and road density, which are consistent with factors influencing the prevalence of COVID-19^[34]^, and with the number of beds per 1,000 people, which are consistent with factors influencing the prevalence of hand, foot, and mouth disease^[30]^, all of which suggest that areas with better economic conditions have a stronger level of health care, higher rates of active medical visits by residents, and higher rates of influenza diagnosis, which may cause better economic areas to have higher influenza incidence rates. However, there is an inconsistency that the influenza incidence rate in Hubei Province is negatively correlated with population density, which may be due to the fact that the deterrent effect of influenza transmission in high-density population areas, such as economic conditions, medical level, and education level, outweighs the promoting effect of population density on influenza transmission, for example, although the population density in urban areas of Wuhan (Jiangan, Jianghan, Qiaokou, and Wuchang districts) is the highest, the influenza incidence rate does not belong to the higher prevalence areas or the highest prevalence areas. In addition, the population base and political area of individual counties also affect the relationship between influenza incidence rate and population density, such as Enshi City, which has a small population base, large political area and relatively low population density, but the influenza incidence rate is as high as 156.96 per 100,000, which is one of the few exceptionally highest prevalence areas in the province. So it seems that the positive effect of socio-economic factors, especially population density on the epidemic of infectious diseases also has a threshold space, and in modern society, not the higher population density is more likely to lead to influenza epidemic.

## 5. Conclusion

Based on the number of influenza incidence cases and incidence rate data in Hubei Province from 2009 to 2019, the spatial and temporal distribution characteristics of the influenza epidemic in Hubei Province and its influencing factors were studied by using time series analysis, ArcGIS spatial analysis, geographical detector, and geographically weighted regression analysis, and the results of this study are as follows:

1. The influenza epidemic in Hubei Province has spatial and temporal characteristics. In terms of temporal variation, the influenza epidemic showed a single peak epidemic in winter and spring during the year, and the overall interannual variation showed a decline followed by an increase, with a tendency to worsen in recent years. In terms of spatial distribution, influenza was generally prevalent but with significant spatial differences, with eastern regions more serious than central and western regions, urban areas more serious than rural areas, with epidemic hot spot areas distributed in the east and west and cold spot areas in the north-central.
2. Influenza epidemic in Hubei Province is the result of the combined effect of natural meteorological factors and socio-economic factors. The seasonal variation of influenza epidemic is mainly influenced by meteorological factors, and the weather of low temperature, low sunshine, low rainfall, high humidity and low wind speed is favorable to influenza epidemic. The interannual variation of influenza epidemic is mainly influenced by socio-economic factors, and the spatial variation of influenza incidence is the result of the interaction between natural meteorological and socio-economic factors, and factors such as road density, number of beds per 1,000 people, urbanization rate and population density have more significant spatial variation on the influenza epidemic.

## Data Availability

Data cannot be shared publicly.

